# Validation, characterization, and utility of markerless motion capture in a large cohort of pediatric patients with complex gait patterns

**DOI:** 10.64898/2026.04.16.26351025

**Authors:** Ross Chafetz, Spencer Warshauer, Sean Waldron, Karen M. Kruger, Seth Donahue, Jeremy P Bauer, Susan Sienko, Anita Bagley, Robert Courter

## Abstract

Markerless motion capture has emerged as a potential substitute for traditional marker-based systems, offering scalable, non-invasive acquisition of human movement. Despite increasing adoption in research and sports applications, its clinical utility for children with complex gait patterns remains an open question. To address this gap, simultaneous marker-based and markerless data were collected in 202 pediatric children (12.1 ± 3.9 years). Marker-based kinematics were processed using the Shriners Children’s Gait Model (SCGM), while markerless outputs were computed using Theia3D with identical Cardan sequences. Agreement between systems was evaluated using statistical parametric mapping (SPM), root-mean-square error (RMSE), and a gait pattern classification based on the plantarflexor–knee extension index. Markerless output systematically underestimated pelvic tilt, hip rotation, and knee rotation and demonstrated reduced between-subject variance in the transverse plane. SPM revealed widespread waveform differences, although most were of negligible effect, especially in the sagittal plane. Mean sagittal-plane RMSEs were < 5° for the knee and ankle and < 8° for the pelvis and hip. Coronal-plane deviations were < 7°, whereas transverse-plane errors exceeded 10°. RMSE increased significantly with body mass index and use of a walker (p < 0.001). Agreement in sagittal-plane gait classification was moderate between systems (κ = 0.60; 67% overall concordance). These results indicate that markerless motion capture is suitable for analyses emphasizing sagittal deviations but remains limited for applications requiring precise axial or frontal-plane estimation. Future work should address algorithmic underestimation of transverse motion and evaluate markerless performance across increasing severity of gait deviation.

## Introduction

Markerless motion capture systems, which use computer vision and deep learning algorithms to estimate human pose without the need for physical markers, offer an appealing alternative to marker based systems (Kanko, Laende, Davis, et al., 2021; Mathis et al., 2018; Wade et al., 2022). These systems are less expensive, have faster setup times, improve patient comfort, and broaden scalability. However, before they can be integrated into clinical practice, their accuracy and reliability must be thoroughly validated against gold-standard marker-based systems. Several studies have examined markerless systems in adults (Antognini et al., 2025; Crespeau et al., 2025; Kanko, Laende, Davis, et al., 2021; Song et al., 2023; Walker et al., 2025) and there are a growing number of studies in pediatric populations, who present unique biomechanical and behavioral challenges (Poomulna et al., 2025; Wishaupt et al., 2024; Wren et al., 2023).

So far, comparisons between marker-based and markerless systems in pediatric populations are relatively limited with regards to sample sizes and scope, but there is good agreement in the sagittal plane and in spatiotemporal parameters (Poomulna et al., 2025; Wren et al., 2023). Perhaps more importantly, markerless systems appear less accurate with more atypical, clinical populations (Poomulna et al., 2025). As motion analysis of gait is considered a standard for surgical decision making in pediatric populations, it is critical that the clinical validity of markerless motion capture be characterized in those with significant deviations.

To provide a clinically meaningful framework beyond statistical measures, we explored the implications of these differences with a classification system developed by Rodda and colleagues for children with spastic diplegia (J. Rodda & Graham, 2001; J. M. Rodda et al., 2004). This schema distinguishes reproducible patterns based on kinematic features of the ankle, knee, hip, and pelvis (J. Rodda & Graham, 2001; J. M. Rodda et al., 2004). It has demonstrated reasonable intra- and inter-rater reliability (Kim et al., 2011; J. M. Rodda et al., 2004), is widely used to guide treatment decision-making (Young et al., 2010), and remains relevant in both clinical and computational approaches to gait analysis (Pantzar-Castilla et al., 2024; Zhang & Ma, 2019). Although originally developed for children with cerebral palsy (CP), we applied this scheme more broadly in our study to use an established, validated framework to test deviations from typical gait and determine whether markerless motion capture yielded classifications consistent with marker-based systems.

The aim of this study was to systematically compare lower-extremity kinematics derived from a traditional marker-based system and a commercially available markerless system using simultaneously collected data in a large pediatric cohort (n = 202) with heterogeneous diagnoses. Agreement was evaluated using complementary metrics, including statistical parametric mapping (SPM) across joint-angle trajectories, root mean square error (RMSE) to classify error magnitude, and sagittal-plane gait classifications to examine concordance in clinically meaningful movement patterns. This represents one of the largest pediatric comparisons of markerless and marker-based gait analysis to date, addressing both quantitative and clinically meaningful agreement between these two approaches.

## Methods

### Procedures

Patients with varying diagnoses underwent clinical gait analysis. Each patient and their guardians (for minors) were provided with informed consent about the study in accordance with the Institutional Review Board Protocol #PHL2305.

Reflective markers were placed on the patient according to the Shriners Children’s Gait Model (SCGM) template (Kruger et al., 2024) by trained and licensed physical therapists. Each patient visit consisted of a static standing trial, a dynamic walking trial to check placement of the knee markers, and a series of dynamic walking trials. All patients completed a barefoot condition while using their necessary ambulation aid, which consisted of – at a minimum – three trials such that three strides could be processed and analyzed.

While the patients ambulated, a marker-based motion capture system comprised of 12 optical, infrared cameras (Vantage V16, Vicon Motion Systems Ltd, Oxford, UK) tracked the reflective markers and 8 FLIR cameras (Blackfly S BFS-U3-23S3C, Teledyne Vision Solutions, Waterloo, ON, Canada) captured 2D digital video for markerless motion capture. Both camera systems were calibrated, synchronized, and captured concurrently through Vicon Nexus (2.15.0) at 60 Hz.

### Data Processing

Marker trajectory data was gap filled and filtered using a 4^th^-order, low pass, Butterworth filter with 10 Hz cutoff. The SCGM was used to calculate joint angles in Python 3.11 (Python Software Foundation, Wilmington, DE, USA).

Digital videos (1920x1200p) from the 8 FLIR cameras were compressed using an H.264 codec before being analyzed in Theia3D (Apollo v2024) (Theia Markerless, Kingston, ON, Canada). Theia computed the transformation matrices which were then used to calculate joint angles in Vicon ProCalc (v1.6.0) using the same Cardan sequences as the SCGM (Kruger et al., 2024). These markerless angles could then be compared to the joint angle outputs from the marker-based SCGM model. Altogether, 12 kinematic variables were estimated bilaterally (i.e., the three-dimensional joint angles about the pelvis, hip, knee, and ankle) from the marker-based and markerless model. Three-to-five trials with complete gait cycles were included per patient. Both the marker-based and markerless outputs were resampled to consist of 101 data points per step.

### Statistical Analysis

Statistical parametric mapping (SPM) provided inferences regarding marker-based and markerless kinematic trajectories while continuous in time (Pataky, 2010; Wishaupt et al., 2024). Average stride kinematics for each participant were utilized, and SPM estimated specific periods during the gait cycle in which differences between marker-based and markerless systems were more apparent using paired t-tests for each joint and plane of motion. To account for multiple comparisons arising from separate SPM analyses across joints and planes, p-values associated with significant clusters were adjusted using the Benjamini–Hochberg false discovery rate correction (*α* = 0.05) (Benjamini & Hochberg, 1995).

To determine whether differences, if any, between systems as determined by SPM were clinically relevant (typically, < 5°), RMSE was first computed separately for each patient by averaging squared differences between the marker-based and markerless kinematic systems across all trials, sides, and time-normalized gait cycle points for each kinematic variable. Group-level RMSE and 95% confidence intervals were then estimated using nonparametric bootstrap resampling at the patient level (5000 iterations). A normalized RMSE was also computed by dividing the RMSE by the total range of motion for each kinematic trajectory across all patients.

Within-subject standardized effect sizes were then calculated using Cohen’s *d* (Cohen, 1988) by computing pointwise differences between systems across the gait cycle, normalizing by the pooled standard deviation at each time point, and averaging the absolute *d* values across time.

Patients were also stratified into multiple groups to investigate factors impacting RMSE. Groupings included age, BMI, sex, and ambulation aid. Age was discretized into three subgroups (3-7, 8-12, and 13+ years). BMI was discretized using established standards (Weir & Jan, 2025); note that all patients considered obese were classified together and not further categorized as Class I-III. Use of an ambulation aid was discretized as either not using an assistive device (“No Assist”) or using an assistive device (“Assist”). To investigate potential effects of these factors on RMSE between systems, a linear mixed effects model was fit to the log-transformed RMSE:

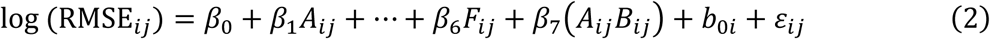

Fixed effects included plane of motion (*A_ij_*) and joint (*B_ij_*) and their interaction, as well as discrete age group, BMI, gender, and assistive device as defined above and in **Table 2**. We log-transformed RMSE to satisfy assumptions of normality and homoscedasticity and placed a random intercept on patient (*i*).

Lastly, using the processed and resampled kinematics, each patient’s gait was then classified algorithmically using the plantarflexor-knee extension index (Sangeux et al., 2015), which is based on the categories defined by Rodda and Graham (J. M. Rodda et al., 2004). For this index, the average z-score within 20-45% of the gait cycle was calculated for each trial’s sagittal knee and ankle kinematics based on a normative distribution computed from our internal, typically developing dataset of 89 patients. Gait was then classified into a category based on the combination of knee and ankle z-scores described in **Table 1**.While the original plantarflexor-knee extension index describes five categories, here we add two additional classifications – recurvatum and ankle crouch (**Table 1**) (Sangeux et al., 2015). While this framework is tailored for CP populations (Kim et al., 2011), its emphasis on sagittal plane deviations from typical during stance makes it applicable across other diagnostic categories.

**Table 1.**
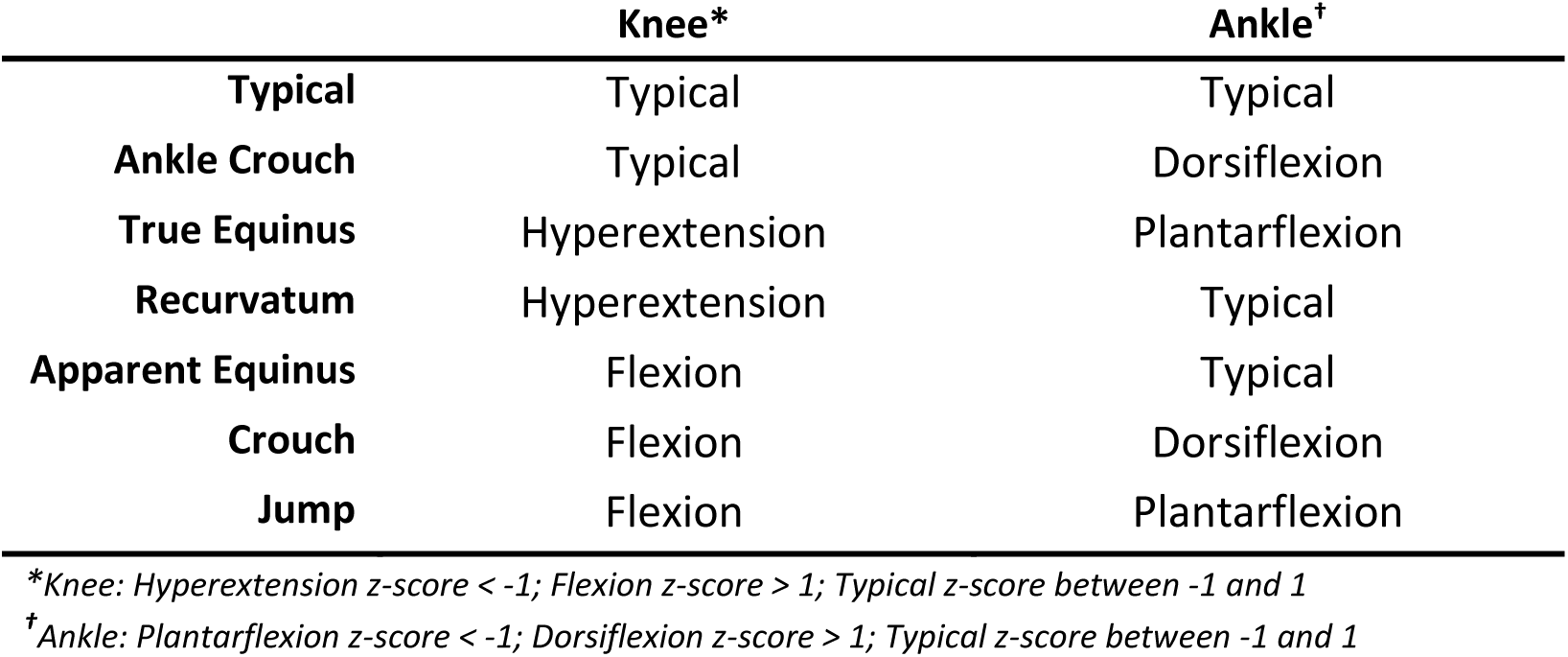
Algorithmic sagittal gait pattern classifications based on the plantarflexor–knee extension couple index.

Agreement of the subsequent classified gait pattern for each trial and side between systems was assessed with an unweighted Cohen’s kappa (*κ*), in which each system was treated as a rater (McHugh, 2012). In total, 1518 steps were classified across all patients. Agreement of ankle and knee plantarflexor-knee extension indices were compared in a similar manner using Cohen’s kappa (*κ*).

## Results

### Patient demographics

An overview of the patient population is reported in **Table 2** (n = 202; 115 males; 12.1 ± 3.9 yrs). Our cohort contained 72 unique diagnoses; however, the most common condition was CP.

**Table 2.** Patient demographics. All values except Mean ± SD are counts.

|  |  | Female<br>(n = 87) | Male<br>(n = 115) |
| --- | --- | --- | --- |
| **Age Group** | Mean ± SD | 12.3 ± 4.0 | 11.9 ± 3.8 |
|  | Age 3-7 | 13 | 22 |
|  | Age 8-12 | 38 | 45 |
|  | Age 13+ | 36 | 48 |
| **BMI** | Mean ± SD | 22.6 ± 7.3 | 20.6 ± 6.0 |
|  | Underweight | 31 | 57 |
|  | Normal | 33 | 35 |
|  | Overweight | 7 | 14 |
|  | Obese | 16 | 9 |
| **Diagnosis** | Cerebral Palsy | 34 | 44 |
|  | Talipes Equinovarus | 4 | 9 |
|  | Arthrogryposis | 6 | 6 |
|  | Spina Bifida | 5 | 5 |
|  | Other | 38 | 51 |
| **Ambulation Aid** | No Assist | 73 | 101 |
|  | Assist | 14 | 14 |

### Kinematics

SPM demonstrated differences throughout a majority of the gait cycle for most kinematic variables, apart from ankle dorsi/plantarflexion and pelvic obliquity (**Fig. 1**). In general, the markerless system tended to underestimate kinematic magnitudes in all planes as compared to marker-based. The most notable differences were found at the hip, knee, and ankle in the frontal and transverse planes. While there were statistical differences throughout the gait cycle at most joints owing in part to the large sample size, the subsequent RMSE calculations provided details as to whether these differences were clinically relevant.

**Figure 1.**
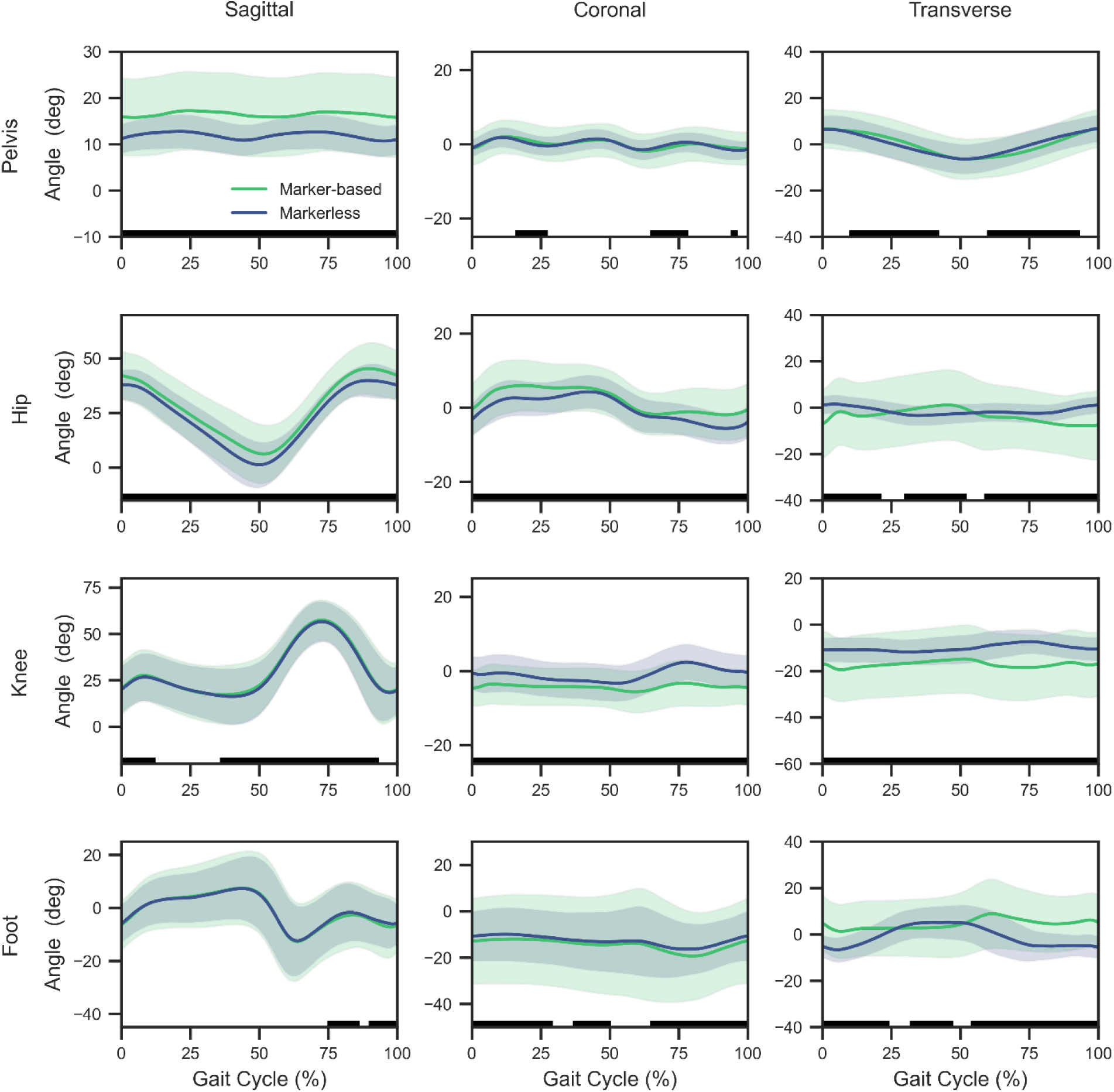
SPM comparing marker-based (green) to markerless (blue) across all patients, trials, and sides. Black bar(s) below graphs indicates regions of statistically significant differences between the systems. Note: the coronal foot displays global foot progression angle, as opposed to inversion/eversion of the ankle.

Bootstrapped RMSE confidence intervals and effect sizes are summarized in **Table 3** for each plane and joint. RMSE between systems was less than 5° for knee flex/extension, ankle dorsi/plantarflexion, pelvic obliquity, hip ab/adduction, and pelvic rotation (**Fig. 2**). Effect sizes for these variables were likewise small (Cohen’s *d* = 0.03-0.37). Pelvic tilt, hip flex/extension, knee varus/valgus, and foot progression angle saw RMSEs between 5 and 10° (Cohen’s *d* = 0.12-0.70). Transverse plane measures at the hip, knee, and ankle were the only kinematic values to exceed 10° RMSE on average, all of which had moderate effect sizes (Cohen’s *d* = 0.34-0.61).

**Figure 2.**
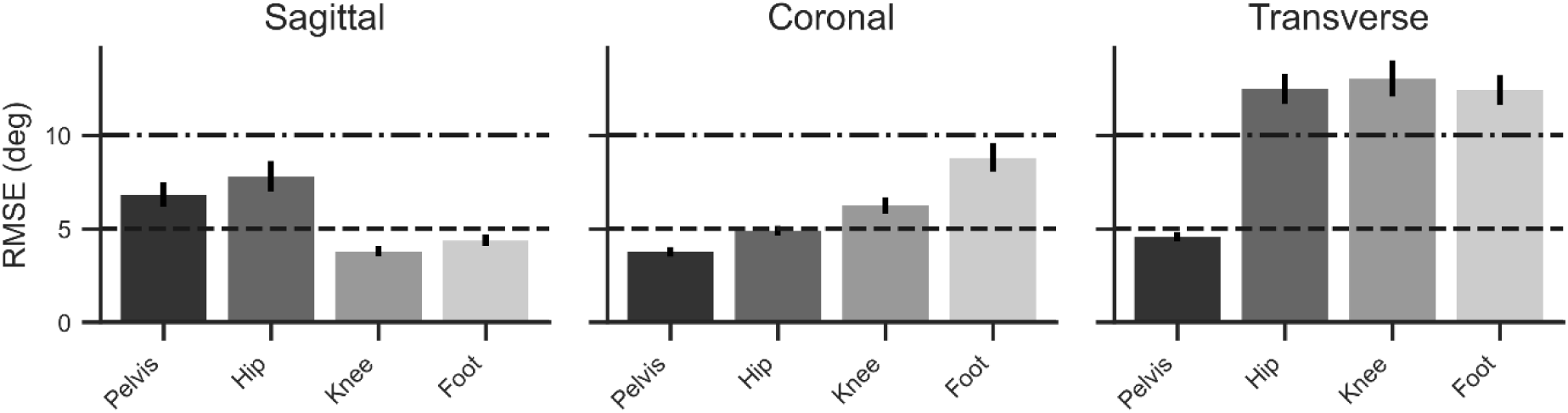
Average RMSE (± 95% bootstrapped confidence intervals) by plane and joint. Note: coronal foot displays global foot progression angle, as opposed to inversion/eversion of the ankle.

**Table 3.** RMSE between systems across all planes and joints.

|  |  | RMSE [95% CI] | Norm. RMSE* | Cohen's *d* |
| --- | --- | --- | --- | --- |
| **Pelvis** | *Sagittal* | 6.81 [6.19, 7.48] | 0.86 | 0.70 |
|  | *Frontal* | 3.77 [3.50, 4.05] | 0.40 | 0.10 |
|  | *Transverse* | 4.57 [4.31, 4.83] | 0.24 | 0.14 |
| **Hip** | *Sagittal* | 7.77 [7.00, 8.59] | 0.18 | 0.42 |
|  | *Frontal* | 4.90 [4.65, 5.14] | 0.32 | 0.37 |
|  | *Transverse* | 12.47 [11.70, 13.29] | 0.60 | 0.34 |
| **Knee** | *Sagittal* | 3.77 [3.51, 4.06] | 0.08 | 0.06 |
|  | *Frontal* | 6.24 [5.82, 6.68] | 0.84 | 0.64 |
|  | *Transverse* | 13.03 [12.04, 14.07] | 0.90 | 0.61 |
| **Ankle** | *Sagittal* | 4.37 [4.05, 4.70] | 0.15 | 0.03 |
|  | *Transverse* | 12.41 [11.63, 13.23] | 0.76 | 0.61 |
| **Foot** | *Transverse* | 8.78 [8.08, 9.54] | 0.39 | 0.12 |

\*Norm. RMSE: normalized RMSE calculated as the RMSE divided by the average range of motion across the gait cycle

Additional covariates also impacted RMSE between systems. RMSE between systems was impacted by BMI, use of an assistive device, and marginally by sex. Overweight (*β* = 0.201 ± 0.056, p < 0.001) and obese individuals (*β* = 0.319 ± 0.053, p < 0.001) saw a 22% and 38% increase in RMSE on average, respectively, as compared to those classified as normal. Underweight individuals did not differ significantly from normal (*β* = 0.032 ± 0.040, p = 0.413). Use of an assistive device – primarily a posterior walker in our sample – increased error by approximately 21% on average compared to those without a walker (*β* = 0.190 ± 0.047, p < 0.001). There was a small effect of gender such that RMSE in males was 7% lower, on average (*β* = 0.070 ± 0.033, p = 0.032). There was no noticeable effect of age group on these system differences (p’s > 0.05).

### Rotational Variation

A Levene’s test indicated that at the hip, variability in rotation from the markerless system was less than that of the marker-based (SD_marker-based_ = 15.1°; SD_markerless_ = 4.6°; p << 0.001) as well as in the frontal plane (SD_marker-based_ = 7.5°; SD_markerless_ = 5.9°; p = 0.002), but not in the sagittal plane (SD_marker-based_ = 18.3°; SD_markerless_ = 15.9°; p = 0.064). At the knee, only rotation saw reduced variance (SD_marker-based_ = 14.6°; SD_markerless_ = 6.0°; p << 0.001), whereas the sagittal and frontal plane motion did not (p = 0.559 and 0.731, respectively).

The lack of variability in the transverse plane along with relatively limited mean hip rotation throughout the gait cycle (**Fig. 1**), suggested that system differences may be exacerbated in those patients with elevated internal or external hip rotation. To investigate, we selected patients with some of the highest degrees of internal and external hip rotation in our cohort and visualized their kinematics. Indeed, patients with large degrees of hip internal or external rotation saw RMSEs around 20-25°, on average, compared to the overall sample’s RMSE of just over 10° (**Table 3**, **Fig. 2**). Qualitatively, the clinical impression of these patients would indicate that some degree of hip external (**Fig. 3**) or internal (**Fig. 4**) rotation should be detected by the motion capture systems; however, in these extreme cases it appears that the marker-based system is better detecting these transverse plane abnormalities of considerable magnitude.

**Figure 3.**
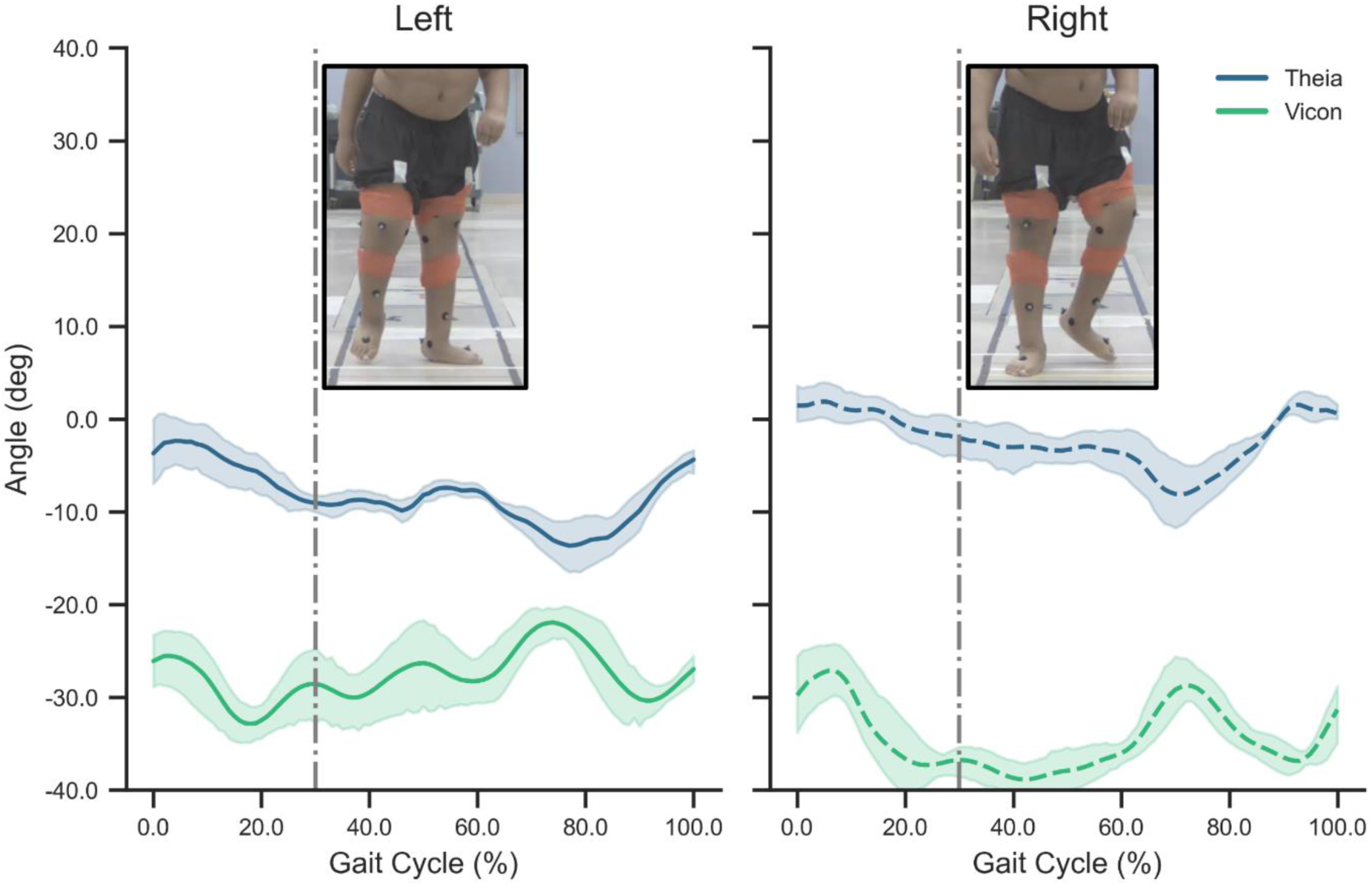
Example of a patient with external hip rotation. Lines and shading are mean ± standard deviation of six trials. The gray line represents the approximate point of interest in the gait cycle corresponding to the photos.

**Figure 4.**
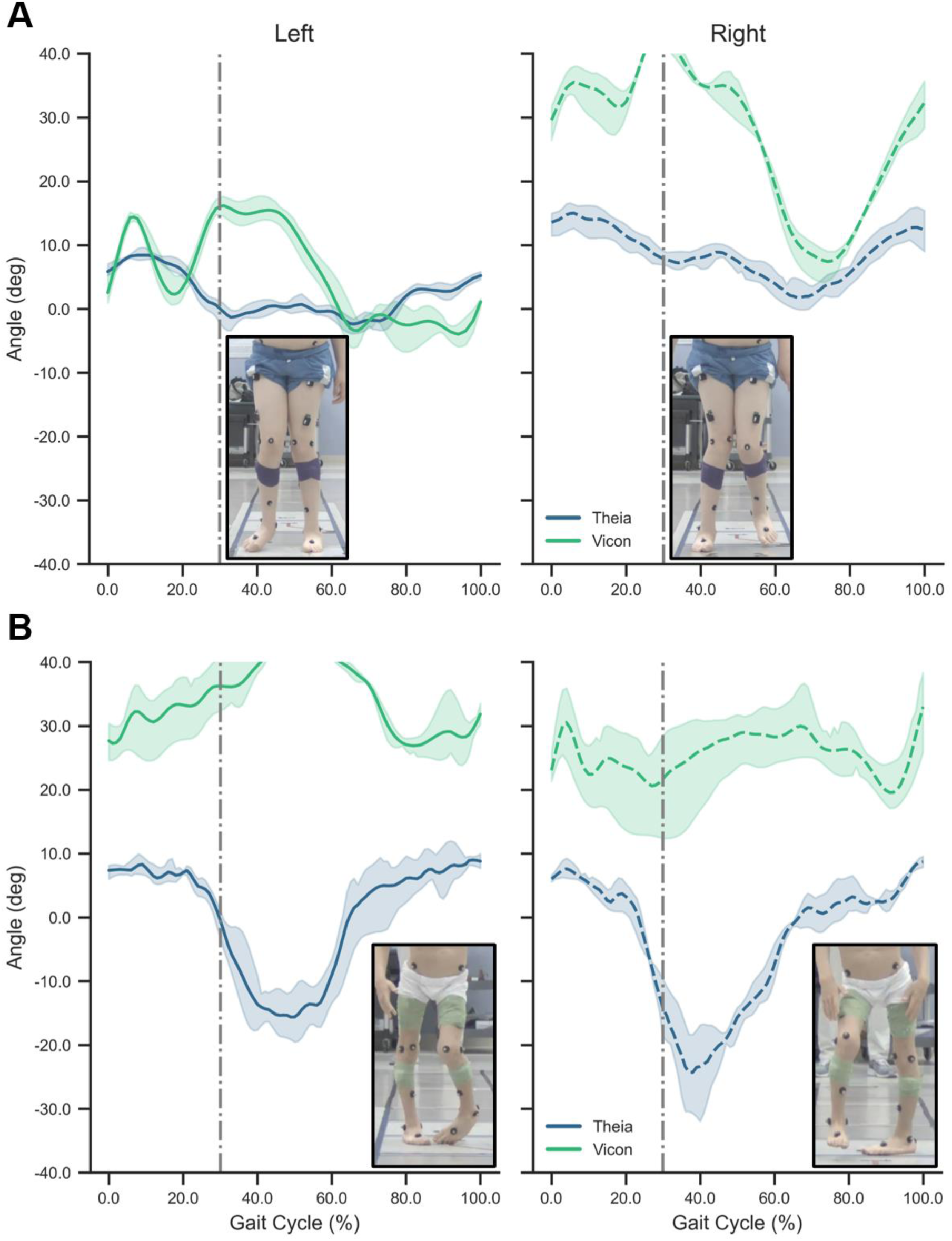
Example of two patients with primarily unilateral (A) and bilateral (B) internal hip rotation. Lines and shading are mean ± standard deviation of three trials. The gray line represents the approximate point of interest in the gait cycle corresponding to the photos.

### Gait classification

We used a gait classification to compare agreement when reducing the multidimensional kinematics down to a single index (J. M. Rodda et al., 2004; Sangeux et al., 2015). Moderate agreement was found in gait classifications when comparing marker-based to markerless classification with κ = 0.60 (**Table 4**). Percent agreement was highest for true equinus (83%), jump (77%), and crouch (69%), and lowest for recurvatum (52%) and ankle crouch (23%).

**Table 4.** Gait classification trial-level agreement and reliability.

| | Category | Trial-Level Agreement | Trial-Level Agreement (%) | Trial-Level Kappa ($\kappa$) |
| --- | --- | --- | --- | --- |
| **Gait Classification** | *Overall* | 1019/1518 | 67% | 0.60 |
|  | *Typical* | 133/225 | 59% |  |
|  | *True Equinus* | 139/168 | 83% |  |
|  | *Jump* | 229/298 | 77% |  |
|  | *Apparent Equinus* | 196/319 | 61% |  |
|  | *Crouch* | 281/409 | 69% |  |
|  | *Ankle Crouch* | 8/35 | 23% |  |
|  | *Recurvatum* | 33/63 | 52% |  |
| **Knee Classification\*** | *Overall* | 1236/1518 | 81% | 0.63 |
|  | *Typical* | 219/332 | 66% | 0.49 |
|  | *Hyperextension* | 116/160 | 73% | 0.65 |
|  | *Flexion* | 901/1026 | 87% | 0.74 |
| **Ankle Classification<sup>†</sup>** | *Overall* | 1199/1518 | 79% | 0.68 |
|  | *Typical* | 491/607 | 81% | 0.58 |
|  | *Plantarflexion* | 403/466 | 86% | 0.79 |
|  | *Dorsiflexion* | 305/445 | 69% | 0.68 |

\*Knee: Hyperextension z-score < -1; Flexed z-score > 1; Typical z-score between -1 and 1
†Ankle: Plantarflexion z-score < -1; Dorsiflexion z-score > 1; Typical z-score between -1 and 1

When viewing the z-scored joint kinematics used in arriving at these classifications, there was strong agreement for the ankle (κ = 0.68; 79% agreement) and the knee (κ = 0.63; 81% agreement) (**Table 4**). Within the knee joint, agreement was highest when detecting flexion (κ = 0.74; 87% agreement), but weakest for typical (κ = 0.49; 66% agreement). At the ankle, agreement was substantial when detecting plantarflexion (κ = 0.79; 86% agreement) and dorsiflexion (κ = 0.68; 69% agreement), but again lowest for typical knee motion (κ = 0.58; 81% agreement). When regressing the marker-based against the markerless z-scores at the knee and ankle, *R^2^* were high at 0.913 and 0.840, respectively (**Fig. 5**).

**Figure 5.**
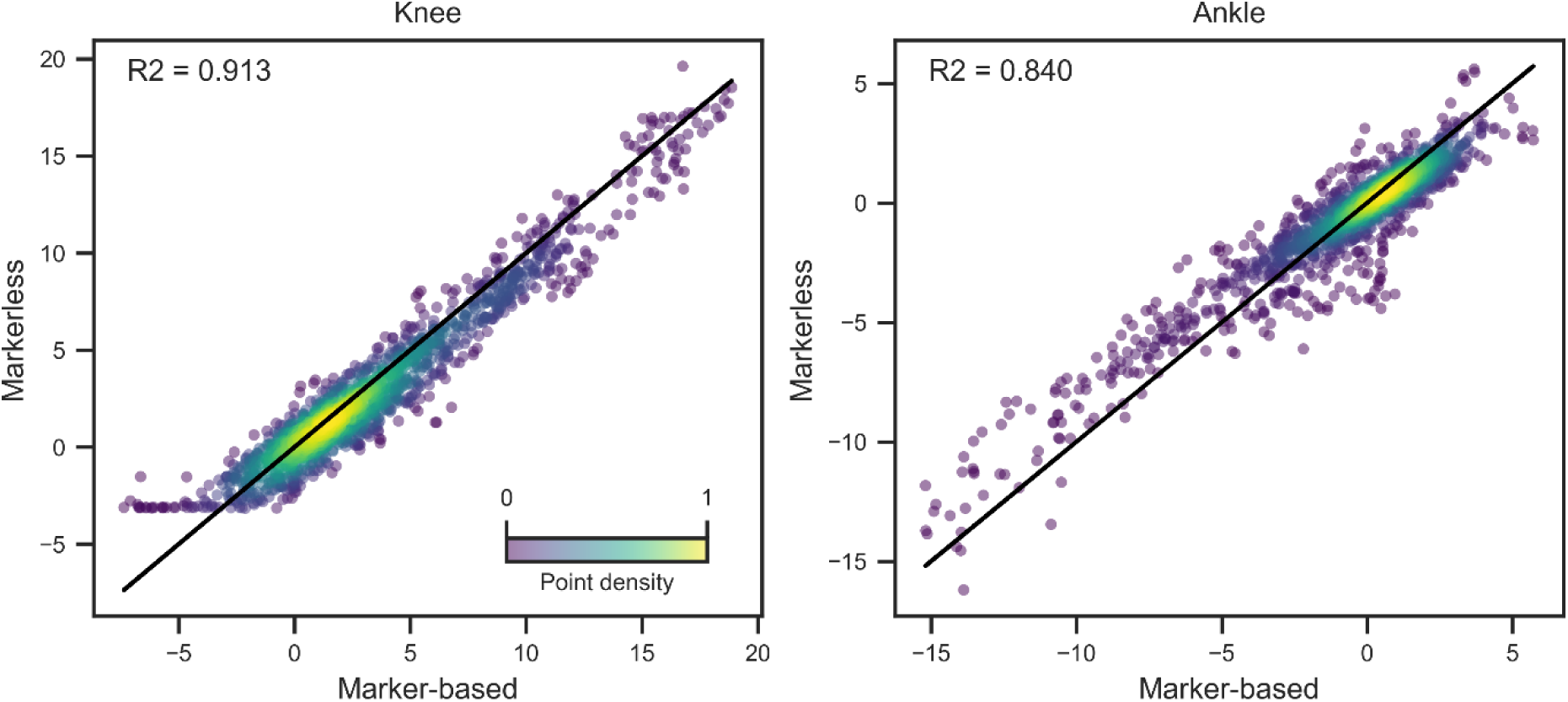
Correlation of Z-scored knee (left) and ankle (right) sagittal kinematics used in the classification process for every stride in the dataset. Note the asymptote of the knee between -5 and 0 is caused by biomechanical constraints of the markerless model preventing excessive knee hyperextension. The solid black line is unity. Color represents the relative density of the data.

## Discussion

The purpose of this study was to evaluate the concurrent validity of markerless motion capture compared to traditional marker-based motion capture in a pediatric population with complex gait patterns. Consistent with prior validation studies in both adults and children, our findings demonstrate good agreement in the sagittal plane and moderate-to-poor agreement in the coronal and transverse planes (Kanko, Laende, Davis, et al., 2021; Poomulna et al., 2025; Song et al., 2023; Walker et al., 2025; Wishaupt et al., 2024; Wren et al., 2023). While this plane-dependent pattern was consistent across covariates, overall error was impacted by sex, BMI, and use of an assistive device. In addition, we found moderate agreement for sagittal-plane gait pattern classification at the knee and ankle (J. M. Rodda et al., 2004; Sangeux et al., 2015), supporting the potential for markerless systems in screening patient’s knee and ankle motion and to contribute to clinically relevant decision-making in certain domains.

### Markerless in the sagittal plane

Sagittal plane performance was the strongest across systems, with RMSE values for the hip, knee, and ankle all below 8°, with knee/ankle errors typically below 5°. While SPM detected differences across the gait cycle at the sagittal knee and hip, in general, these differences were minor compared to the total range of motion at those joints and had small to moderate effect sizes, respectively. The sagittal ankle had some of the strongest agreement, with low RMSE and temporal differences only during late swing. However, at the pelvis, markerless consistently estimated less pelvic tilt than the marker-based system. This may be attributed to differences in pelvis segment definitions across the two biomechanical models, and subsequent underestimation of hip flexion is likely a downstream effect of these pelvic orientation differences. These discrepancies could be addressed through offset adjustments or by developing markerless-specific normative datasets to avoid systematic bias (Antognini et al., 2025; Kanko, Laende, Davis, et al., 2021)

When classifying gait algorithmically (J. M. Rodda et al., 2004; Sangeux et al., 2015), agreement between systems was moderate across all knee and ankle categories, suggesting clinical utility for detecting deviations relevant to intervention planning. This is particularly notable considering recent work advocating for motion analysis in the early detection of knee deformity to guide less invasive interventions (Thomason et al., 2025).

The lowest trial-level agreement was observed for ankle crouch, recurvatum, apparent equinus, and typical patterns. These classifications involve one or both joints being labeled as “typical” (−1 ≤ z ≤ 1), in which these tighter bounds may make them sensitive to small kinematic differences that shift a joint to a different classification. In contrast, for example, true equinus showed high agreement, as both knee and ankle criteria are unbounded in one direction (knee hyperextension and ankle plantarflexion; z < −1). Here we made classifications based on a normative dataset collected with marker-based systems; a potential solution to improve classification accuracy would be to calculate the markerless z-scores relative to a normative markerless dataset.

### Coronal and transverse plane performance

Coronal and transverse plane performance, however, was less robust. In marker-based systems, transverse knee kinematics are known to be susceptible to error due to examiner variability in marker placement (Baker et al., 1999; Jensen et al., 2016) and soft tissue artifacts (Wade et al., 2022). This can lead to crosstalk between rotational and varus/valgus signals, particularly during swing phase. While changes such as eliminating the use of a thigh wand for a patella marker and careful alignment checks have improved transverse plane accuracy in marker-based systems, variability remains (Baker et al., 1999; Jensen et al., 2016; McMulkin & Gordon, 2009; Veilleux et al., 2025).

Markerless systems offer a theoretical advantage by eliminating examiner-induced variability (Kanko, Laende, Selbie, et al., 2021). However, it appeared that a potential shortcoming of markerless may be a simultaneous reduction in the between-subject variability it currently detects. For example, the marker-based data exhibited substantial between-subject variability in hip rotation, which is consistent with the heterogeneity of our clinical cohort; in contrast, markerless outputs showed markedly reduced variance with waveforms clustered near neutral throughout the gait cycle (Wishaupt et al., 2024). This divergence is likely attributable to algorithmic and modeling effects rather than a true physiological finding. Video-based estimation of axial rotations is intrinsically challenging, and it is not clear if the training set contained a sufficiently diverse population to identify atypical hip and knee transverse plane motion (Kanko, Laende, Davis, et al., 2021; Needham et al., 2021). The persistence of normal variability in sagittal-plane measures alongside suppressed transverse-plane variability supports a rotation-specific limitation.

To further examine these discrepancies, we analyzed a subset of children with extreme hip internal or external rotation in the marker-based data. Visual inspection of gait videos confirmed pronounced transverse-plane deviations that were accurately reflected by marker-based kinematics, whereas markerless output consistently underestimated these magnitudes or showed opposing trends (e.g., **Fig. 4**). These results indicate that markerless motion capture does not reliably quantify extreme hip rotation, and transverse-plane findings should therefore be interpreted with caution in clinical populations when accurate rotational assessment is required for treatment decisions.

### Strengths and limitations

This study leveraged a large, heterogeneous pediatric cohort (>200 patients) spanning a wide range of ages, body sizes, and gait patterns, strengthening the generalizability of the findings. Future work will further stratify analyses by diagnosis and gait severity, incorporate markerless normative datasets, and evaluate additional clinically meaningful outcomes (e.g., GDI, GPS, EVGS). Given observed limitations in rotational kinematics, characterizing markerless performance as a function of gait deviation severity will be particularly important.

Despite the sample size, all data were collected in a single laboratory, limiting assessment of environmental factors such as lighting and camera configuration on markerless accuracy. Ongoing multi-site data collection will allow evaluation of markerless feasibility across varied clinical settings. Accuracy was influenced by intrinsic patient factors, including assistive device use (Wren et al., 2023), BMI, and sex, with higher RMSE observed in patients using assistive devices and those with obesity. However, because both marker-based and markerless systems are affected by increased soft tissue and modeling assumptions, relative accuracy remains unclear (Leardini et al., 2005).

Finally, it is important to recognize that marker-based models also rely on assumptions that impact kinematic estimates, particularly hip joint center localization (Kainz et al., 2015). Common predictive approaches, such as the Harrington equations used in the SCGM (Harrington et al., 2007; Kruger et al., 2024), are based on limited pediatric data and can substantially influence downstream joint angles (Stagni et al., 2000; Zuk et al., 2014). Continued efforts to improve hip joint center estimation and to expand markerless model training using atypical pediatric gait data are therefore critical, regardless of motion capture modality.

## Conclusion

In summary, this study demonstrates that markerless motion capture provides clinically useful estimates of sagittal-plane kinematics in a pediatric population, with good agreement to marker-based systems and potential for integration into clinical workflows. The system offers substantial efficiency gains and reduced patient burden, making it particularly attractive for pediatric settings. However, its performance in the coronal and transverse planes was notably weaker, with systematic underestimation of hip rotation and suppression of expected inter-subject variability. These limitations indicate that transverse-plane outputs should be interpreted with caution and are not currently suitable for clinical decision-making. Thus, while Theia may be adopted with reasonable confidence for sagittal-plane analyses, further refinement of underlying models and continued validation in diverse patient populations will be necessary before markerless kinematics can reliably replace marker-based methods in the transverse domain.

## Data Availability

All data produced in the present study are available upon reasonable request to the authors

## Acknowledgements

We would like to acknowledge the physical therapists (PT, DPT), namely Jacqueline Weiss, Emma Riegert, and Mallory Meyer for their roles in data collection. We have no funding sources to acknowledge.

